# Provision of a liquefied petroleum gas cookstove and fuel during pregnancy and infancy and linear growth trajectories between birth and 12 months: evidence from the multi-center Household Air Pollution Intervention Network (HAPIN) trial

**DOI:** 10.1101/2025.06.05.25329099

**Authors:** Kasthuri Sivalogan, Aryeh D. Stein, Lisa M. Thompson, Jiantong Wang, Anaite Diaz-Artiga, Vigneswari Aravindalochanan, Shirin Jabbarzadeh, Laura Nicolaou, Kendra N. Williams, Kalpana Balakrishnan, Jennifer L. Peel, William Checkley, Thomas Clasen, Sheela S. Sinharoy, HAPIN investigators

## Abstract

**Objective:** Exposure to particulate pollution from cooking with solid biomass fuels is associated with impaired child linear growth. We examined the effect of a liquefied petroleum gas (LPG) cookstove randomized control trial during pregnancy and infancy on linear growth trajectories among infants born to women enrolled during pregnancy.

**Methods:** The Household Air Pollution Intervention Network (HAPIN) randomized control trial enrolled 3195 pregnant women (9 to <20 weeks gestation) from rural areas in Guatemala, Peru, India, and Rwanda that relied primarily on biomass fuels for cooking. Women in the intervention group received an LPG cookstove and fuel for approximately eighteen months, while those in the control group continued to use biomass for cooking. We measured the children’s recumbent length at birth and 3, 6, 9 and 12 months of age and calculated length-for-age z-score (LAZ). We conducted a multiple group latent class growth analysis among the 2802 infants who finished the study and had ≥3 length measurements across the five timepoints to examine if latent classes differed by intervention arm.

**Results:** We identified three latent classes of linear growth, based on visual inspection of mean LAZ and model fit statistics, that represent higher, medium, and lower LAZ trajectories. Approximately 13.2% of infants belong to the high LAZ trajectory, 53.8% of infants belong to the medium LAZ trajectory and 33.0% belong to the low LAZ trajectory. The distribution of infants in each latent class did not differ by intervention assignment.

**Conclusions:** Provision of an LPG cookstove and fuel during pregnancy and infancy did not alter linear growth trajectories among the offspring.

**Clinical Trials Registration Number:** NCT02944682

**Funding Source:** U.S. National Institutes of Health and the Bill & Melinda Gates Foundation

## INTRODUCTION

Household air pollution, including from the incomplete combustion of solid fuels for cooking and heating using poorly ventilated combustion devices, continues to be a leading risk factor for global morbidity and mortality [1]. Women bear primary responsibility for cooking at the household level and therefore are at higher risk of cooking-related household air pollution exposure relative to other household members [2]. Women are also often primary caregivers for young children, and children’s physical proximity to their mothers during sensitive periods of growth and development between birth and two years of age may increase their own risk of household air pollution exposure [2, 3]. Air pollution exposure is one of the leading threats to children’s health, accounting for almost 1 in 10 deaths in children under-five [4].

Prenatal household air pollution exposure can impair linear growth via oxidative stress and systemic inflammation, while postnatal exposure can result in impaired immune development and function, clinical and subclinical infection, dietary intake and metabolism and bone metabolism [3]. Risk of impaired linear growth is also associated with poor child development and increased risk for all-cause mortality, infectious disease mortality and chronic disease development in adulthood [3, 5].

However, most of the evidence to date on household air pollution exposure and linear growth is derived from observational studies that report associations between estimated exposure at birth and growth outcomes [6]. A systematic review and meta-analysis of 11 studies with 168,298 children concluded that there was a 19% (95% CI: 1.10, 1.29) increased risk of stunting associated with exposure to household air pollution among children under five [7]. Most studies estimated household air pollution exposure based on self-reported fuel type and complementary determinants, including stove type, household ventilation and cooking habits (location, duration or frequency) without directly measuring exposure; furthermore, most studies were cross-sectional with only one being a prospective cohort [7]. In addition, there is no multi-country evidence of household air pollution exposure on longitudinal growth patterns in the literature [8, 9]. Additional evidence from longitudinal analyses is necessary to capture the rapid changes in growth that occur during early childhood.

The Household Air Pollution Intervention Network (HAPIN) study is a randomized control trial, designed to compare the effects of provision of a liquefied petroleum gas (LPG) cookstove and fuel to continued use of solid fuels. The trial had three primary outcomes focused on infants: low birth weight, severe pneumonia, and stunting at 12 months. Birth weight of infants did not differ significantly between infants born to women in the intervention arm, compared to those in the control arm [10]. Similarly, infants born to women in the intervention arm and infants born to women in the control arm had a similar mean length-for-age z-score (LAZ) at 12 months [8].

Here we report on linear growth trajectories between birth and 12 months for the intervention and control arms of the HAPIN trial. The aim of this analysis is to describe longitudinal changes in LAZ among children 0-12 months participating in the trial. Specifically, we examine patterns of growth by intervention arm, country site, child sex, and timing of intervention delivery.

## METHODS

### Trial design and setting

HAPIN is a multi-center, parallel-group individual randomized control trial (RCT) to assess the effects of an LPG cookstove and fuel intervention on four primary outcomes: birth weight, stunting, and severe pneumonia in infants, and systolic blood pressure among older adult women (Clasen et al. EHP 2020).

HAPIN investigators aimed to enroll 3200 pregnant women across four rural sites: Jalapa, Guatemala; Tamil Nadu, India; Puno, Peru; and Kayonza, Rwanda. Pregnant women were identified from clinical registries, prenatal clinics and/or referred by community health workers. Women were eligible to participate if they were ages 18 to <35 years, cooked primarily with biomass stoves, lived in the trial area and were pregnant with a confirmed singleton fetus between gestational weeks 9 and 20. Following written informed consent, baseline surveys and assessments, pregnant women and their households were randomized to the intervention or control groups. Intervention households received a free LPG stove and LPG delivered to their household as needed for approximately 18 months (for the remainder of the pregnancy and through age 12 months of the child), while control households continued to use their traditional cooking fuel (principally wood, charcoal, and dung). Additional details of the intervention design have been described elsewhere [11].

The study protocol was reviewed and approved by institutional review boards or ethics committees at Emory University (00089799), Johns Hopkins University (00007403), Sri Ramachandra Institute of Higher Education and Research (IEC-N1/16/JUL/54/49), the Indian Council of Medical Research – Health Ministry Screening Committee (5/8/4-30/(Env)/Indo-US/2016-NCD-I), Universidad del Valle de Guatemala (146-08-2016/11-2016), Guatemalan Ministry of Health National Ethics Committee (11-2016), A.B. PRISMA (CE3571·16), the London School of Hygiene and Tropical Medicine (11664-5), the Rwandan National Ethics Committee (No.357/RNEC/2018), and Washington University in St. Louis (201611159). Participants provided written informed consent. The trial is registered with ClinicalTrials.gov (Identifier NCT02944682; URL: https://clinicaltrials.gov/study/NCT02944682). The trial protocol (including changes, approach to harms, stopping guidelines, and planned primary and secondary outcomes) and statistical analysis plan can be accessed at https://clinicaltrials.gov/study/NCT02944682. De-identified data, statistical code and other materials can be accessed upon request.

### Randomization and Masking

Participants were randomly assigned in a 1:1 ratio stratified by setting (ten geographic strata, as listed above) in permuted blocks of two and four to either receive the intervention or continue their traditional cooking practices with biomass fuels. We sought to randomly assign 1600 participants to the intervention arm and 1600 to the control arm. Only one pregnant woman per household was allowed to participate. The HAPIN Data Management Core generated randomization lists based on block randomization using randomly selected block sizes, then prepared individual sealed envelopes containing trial group allocation, which were shipped to each study site and selected by participants. Due to the nature of the intervention, it was not possible to mask participants or data collection teams to the group assignment. However, the study investigators were blinded to the collected data, and primary analyses were conducted on blinded data. Participants in the control group received compensation, which varied by country, to offset the economic benefit of the intervention [12].

### Household Air Pollution Exposure

Personal particulate matter less than 2.5 micrometers in diameter (PM_2.5_), black carbon (BC) and carbon monoxide (CO) exposure was measured three times during pregnancy –at baseline (9 to <20 weeks gestation), between 24-28 weeks gestation and between 32-36 weeks gestation [13]. Full details of the methods to assess personal exposure were previously described [13–15].

### Intervention Fidelity and Adherence; Exposure Contrast between Intervention and Control

There was high fidelity and adherence to the LPG intervention in the HAPIN trial [15, 16]. In addition, there were large exposure contrasts in PM_2.5_, BC and CO between intervention and control arms at the first and second follow-up visits [14]. In the post-intervention follow-up period during pregnancy, 69% of intervention-group PM_2.5_ exposures were below the World Health Organization Annual Interim Target 1 compared to 23% of control-group PM_2.5_ exposures [14] indicating that the LPG intervention successfully reduced household air pollution exposures [14].

### Anthropometric Outcomes

Recumbent length was measured twice to the nearest 1 mm with a Seca 417 measuring board (Seca, Hamburg, Germany) during household visits at five time points: birth (measured in hospital within 24 hours of delivery) and 3, 6, 9 and 12 months of age (+/- 14 days). If the two length measurements differed by >0.7 cm, a third measurement was taken [17].

The COVID-19 pandemic resulted in some interruptions to data collection, and specifically to collection of anthropometric measurements. However, there were no differences in the proportion of children with a missing length measurement between intervention and control arms before or after March 17, 2020 [8].

### Data Management

Recumbent length was calculated as the average of the two closest measurements and converted to LAZ using the WHO Child Growth Standards SAS igrowup package [18]. Participants were included in the analysis if they had ≥3 length measurements across the five time points, to improve model stability [19]. Patterns of missing data by country site, timepoint, intervention arm and infant sex are provided in the Supplemental Materials (**Table #S1**). Missing data are a minimal concern when missing at random in latent class models [20].

### Covariates

Maternal age, maternal education and maternal dietary diversity were assessed at baseline. Maternal education was categorized as 1) no formal education or some primary school, 2) primary school or some secondary school or 3) secondary, vocational or some university. Maternal dietary diversity was assessed using the tool adapted from the Food and Agriculture Organization of the United States to calculate Minimum Diet Diversity for Women [21]. Women were asked to recall the frequency of consumption of a prespecified list of food groups during the last 30 days at baseline and again at 6 months post-partum that were then categorized into ten categories: grains, white roots, tubers, and plantains; pulses; nuts and seeds; dairy; meat, poultry, and fish; eggs; dark green leafy vegetables; other vitamin A-rich fruits and vegetables; other vegetables; and other fruits. Individual consumption was summed into a score ranging from 0-10 based on yes/no responses to the prespecified food groups and each woman was classified as achieving dietary diversity using a cut-off value of ≥5 [22]. Household food insecurity was assessed at baseline and classified according to the Food Insecurity Experience Scale [23]. Socioeconomic status was assessed at baseline and calculated as a derived index that incorporates water and sanitation quality, access to electricity, number of people in the household, food insecurity, participant’s education level, and type of floor, wall and roofing material.

### Statistical Analysis

We used a multiple group latent class growth analysis to examine if patterns of linear growth from birth to 12 months of age across the HAPIN sites vary as a function of intervention arm (Model 1), controlling for the fixed effects of randomization strata [24]. Latent class growth modeling (LCGM) is a longitudinal analytical technique based on structural equational modeling that examines how individuals change by an observable outcome variable over time. In traditional growth modeling, it is assumed that all individuals in the study sample come from a single population and have one average trajectory to describe the underlying pattern of that single population. Individual differences are then captured by random slopes and random intercepts. In LCGM, individuals derive from multiple latent (or underlying) classes to identify distinct patterns within the study population. LCGM is a suitable approach for modeling repeated and highly correlated measures, such as early-life anthropometric measurements, and has a lower computational burden compared to growth mixture modeling [25, 26]. The multiple group analysis, using the “known class” function facilitates use of a grouping option to identity the variable in the dataset that contains information on group membership [27]. All models are adjusted for missing data under the assumption that data are missing at random using the full information maximum-likelihood (FIML) approach [28].

First, we plotted the trajectory of the mean LAZ score to verify longitudinal change and to identify the most appropriate pattern of change. Two measurements are needed to identify a linear model while three or more measurements are needed to identify a quadratic model. Then, we used individual LAZ scores at the five time points to identify the appropriate number of latent classes determined by model fit statistics, model parsimony and interpretability [26]. We used time scores (0-4) to reflect the five timepoints in our model because the timepoints were equidistant and to keep the quadratic time score smaller. We used Bayesian Information Criteria (BIC) to assess model fit and the Bootstrap Likelihood Ratio (BLRT) and Lo, Mendell, and Rubin Likelihood Ratio tests (LMR-LRT) to determine the number of latent classes [26]. We used entropy values to examine the separation of latent classes and the accuracy of classification into specific classes. Finally, we examined the posterior probabilities of an observation classified in each class to examine the percentage of the sample in each class [29]. We selected the final number of latent classes based on BIC, entropy, ability to make comparison across different models, and not having small cell sizes in each class (<5%).

We also examined descriptive statistics by intervention arm and latent class trajectory group to assess if there were differences in sociodemographic characteristics by class membership.

All analyses were conducted in Mplus version 8[30] and R version 4.2.2[31].

### Subgroup Analyses

We examined latent class growth trajectories separately by HAPIN country (Model 2), infant sex (Model 3), and timing of the intervention delivery (Model 4) within the pooled sample. The multiple group analytical method allows class membership and model terms to vary by the variable of interest (e.g., by HAPIN country site) within each model. All subgroup analyses were prespecified. Due to the heterogeneous country settings, sub-analyses by country site are standard for the HAPIN trial. Additionally, there is evidence in the literature for heterogenous impact of household air pollution exposure on infant length by sex [9, 32]. We also examined LAZ growth trajectories by sex separately for each country site (Models 5-8). Prior analysis of the HAPIN intervention effect on birth weight suggested that infants of mothers who received the intervention prior to 18 weeks gestation had slightly higher birth weight than infants of mothers who received the intervention at 18 weeks or later [10]. Therefore, we examined whether LAZ growth trajectories differed by timing of the intervention (<18 weeks compared to ≥18 weeks) (Model 4).

We also controlled for the fixed effects of the randomization strata (trial sites within each country) in models examining latent class trajectories by infant sex and timing of intervention delivery in the pooled sample.

## RESULTS

### Study Population and Participant Characteristics

Between May 7, 2018, and February 29, 2020, 3200 pregnant women were randomized. Five participants were found to be ineligible after randomization, resulting in 3195 pregnant women participating in the trial. In addition, 4.2% (n=134) exited the trial before birth of the child and another 3.5% (n=112) exited the trial after the birth of the child but before 12 months of age. Reasons for withdrawal or exiting the trial are provided in the Supplemental Materials (Table #S2). Harms and unintended events are described in Checkley et al., 2024 [8]. We excluded participants with two or fewer LAZ measurements across the five timepoints (n=147), as indicated in Supplemental Figure #1. The final sample for this analysis included n=2802 children.

Descriptive characteristics for the analytical sample and by intervention assignment are presented in Table 1. Descriptive characteristics by HAPIN country site are provided in the Supplemental Materials (Table #S3). Overall, women were enrolled in the HAPIN trial at 15 weeks gestation and reported consuming an average of three food groups on a daily basis. Women in both the intervention and control groups were of similar height and age. A higher percentage of women in the control group were nulliparous compared to women in the intervention group.

**Table 1.** Descriptive statistics by intervention assignment for 2802 mothers of children that completed participation in the HAPIN Trial.

|  | **Overall**<br>**(n=2802)** | **Intervention**<br>**(n=1412)** | **Control**<br>**(n=1390)** |
| --- | --- | --- | --- |
| **Maternal height at baseline -Mean (SD)** | 152.19 (6.18) | 152.24 (6.29) | 152.13 (6.08) |
| **Maternal age at baseline (Mean, SD)** | 25.41 (4.47) | 25.34 (4.37) | 25.48 (4.56) |
| **Gestational age at baseline (Mean, SD)** | 15.39 (3.11) | 15.52 (3.06) | 15.26 (3.16) |
| **Nulliparous % (N)** | 37.55 (1050) | 35.52 (493) | 39.56 (557) |
| **Socioeconomic index (Mean, SD)** | -0.01 (1.05) | 0.05 (1.02) | -0.08 (1.07) |
| **Minimum dietary diversity (Mean, SD)** | 3.41 (1.57) | 3.46 (1.61) | 3.37 (1.54) |
| **Household food insecurity (Mean, SD)** | 1.38 (2.09) | 1.28 (2.05) | 1.48 (2.13) |

### LAZ Latent Class Trajectories by ITT

Mean LAZ at birth, 3, 6, 9, and 12 months are presented in Table 2, and model fit statistics are presented in the Supplemental Materials (Table #S4). The log-likelihood and BIC decreased as the number of classes increased from a one to four class model (Table #S4). Entropy was >0.80, indicating that latent classes are highly discriminating only for the three and four class models [33]. We determined that a three-class quadratic pattern of change best fit the LAZ data, based on visual inspection of mean LAZ and after evaluating the model fit statistics. We identified three LAZ latent class trajectories for both the intervention and control arms, as indicated in Figure 2.

**Figure 1.**
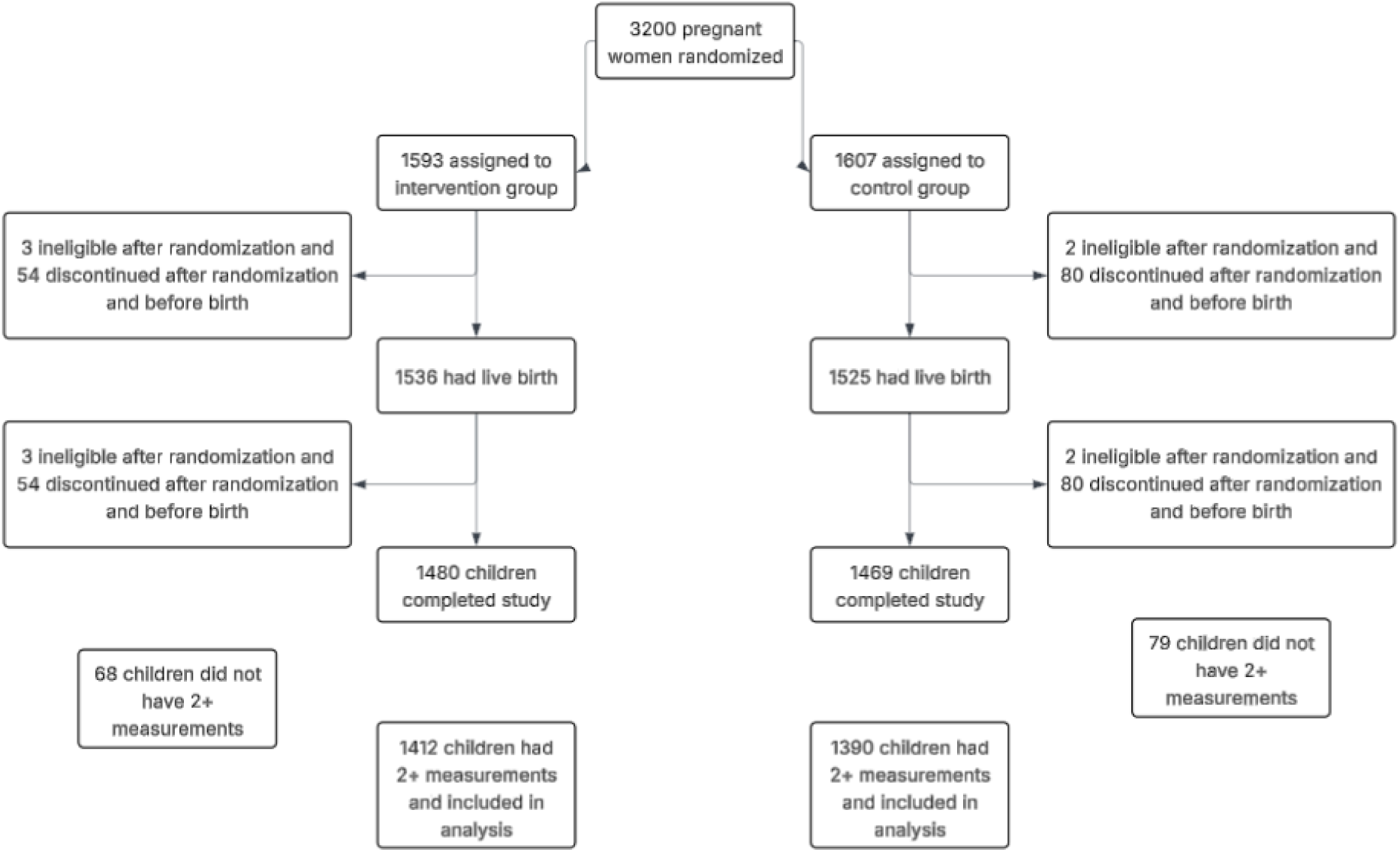
Final sample for analysis

**Figure 2.**
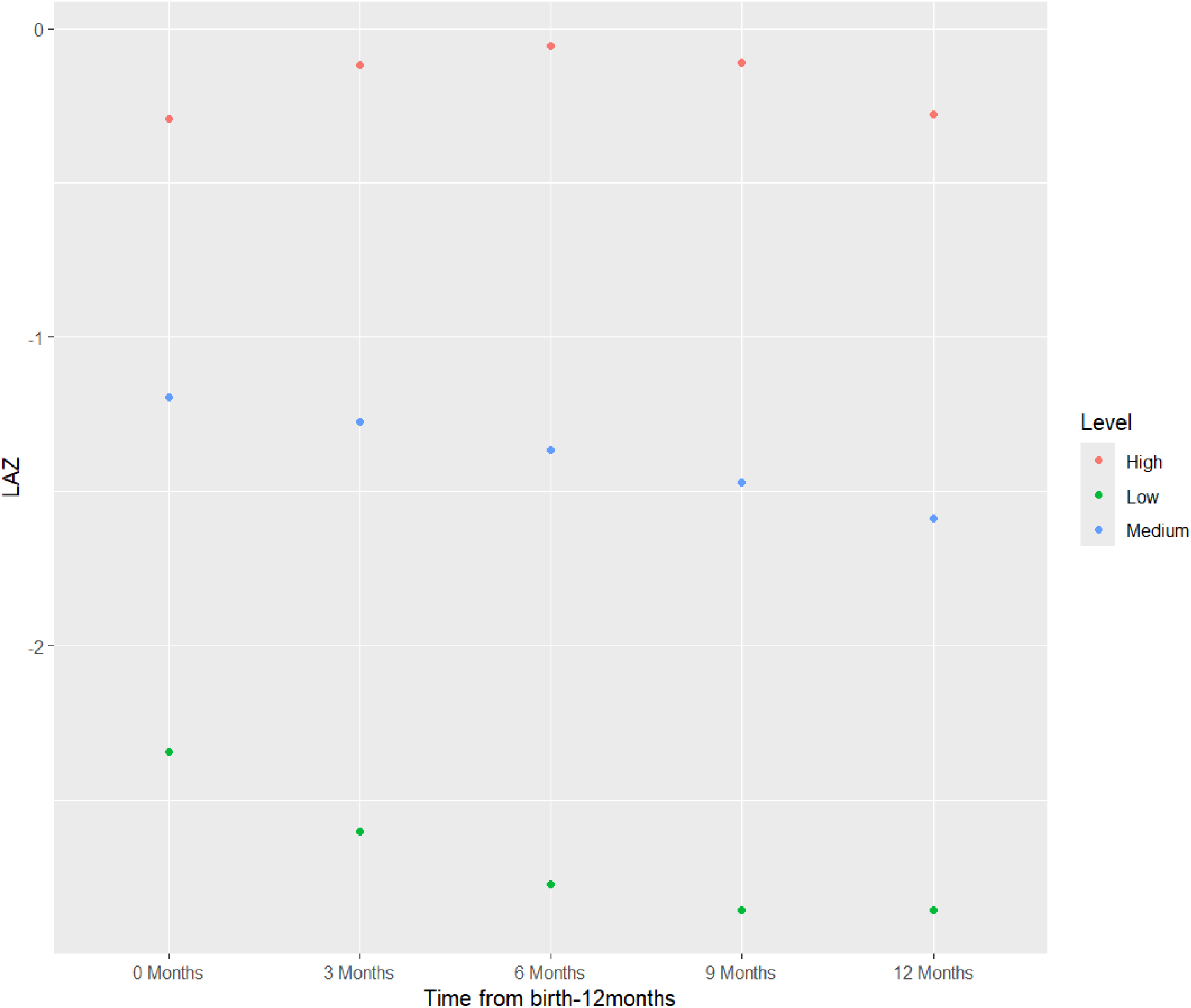
Latent classes for intention-to-treat analysis

**Table 2.**
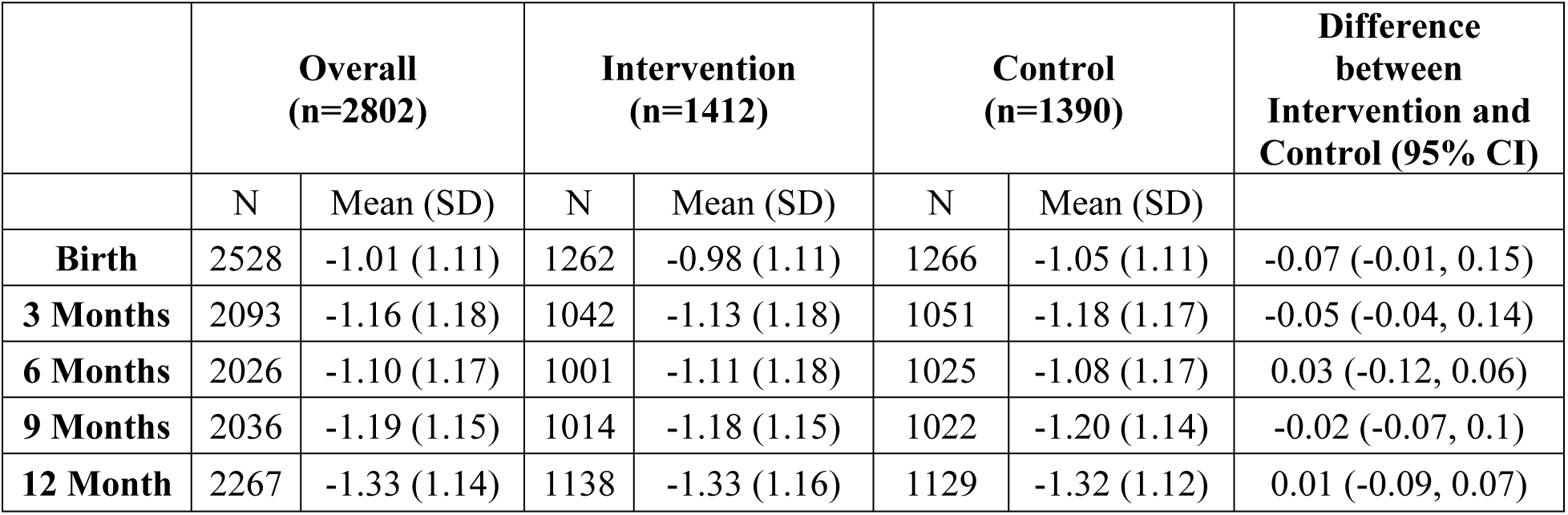
Observed mean LAZ from birth-12 months of age for 2802 children with at least three measurements.

The three distinct latent class trajectory groups, with unique intercepts and slopes, are labelled as “high”, “medium” and “low”. The percentages of infants in each latent class were not significantly different between intervention (higher: 12.3% of infants, medium: 27.3%, lower: 10.8%) and control (higher: 10.6%, medium: 27.9%, lower: 12.1%) arms (p>0.9). There was no overlap in intercepts or slopes between classes for either intervention or control groups. In addition, there were no significant differences in latent class LAZ growth trajectories between the intervention and control groups (p>0.9) (Figure 2). For all three classes, the intervention group had a higher, but not significant, intercept and mean LAZ at all five timepoints as compared to the control group. Members of the high trajectory group in both intervention and control arms exhibited an increase in mean LAZ from birth to 6 months followed by a decrease from 6 to 12 months. Members of the medium and low trajectories in both intervention and control arms exhibited an overall decrease between birth and 12 months. Additionally, members of the low trajectory in both intervention and control arms had a mean LAZ less than 2 standard deviations (LAZ < −2 SD) from the mean from birth through 12 months of age.

We examined sociodemographic characteristics by intervention assignment and latent class trajectory group (Table 3). In the intervention arm, there were differences in household socioeconomic index, maternal minimum dietary diversity score, maternal height, preterm births (birth that occurs before 37 weeks gestation), and the percentage of female infants born to mothers by latent class trajectories. In the control arm, there were additional differences in household food insecurity score and the percentage of female infants born to mothers.

**Table 3.** Descriptive statistics of study participants by LAZ latent class trajectories for 2802 children with at least three measurements and whose mothers completed the HAPIN Trial.

|  | Trajectory Group |  |  |
| --- | --- | --- | --- |
|  | High<br>(N=369.45) | Medium<br>(N=1506.86) | Low<br>(N=925.68) |
| **Maternal height at baseline (Mean, SD)** | 149.34 (6.11) | 151.69 (5.93) | 154.09 (6.07) |
| **Maternal age at baseline (Mean, SD)** | 25.32 (4.61) | 25.33 (4.50) | 25.59 (4.35) |
| **Gestational age at baseline (Mean, SD)** | 15.51 (3.13) | 15.29 (3.08) | 15.51 (3.15) |
| **Nulliparity (Yes) (N, %)** | 117 (34.11) | 580 (37.76) | 353 (38.50) |
| **Infant sex (Female) (N, %)** | 116 (33.72) | 746 (48.50) | 492 (53.48) |
| **Preterm birth (Yes) (N, %)** | 56 (16.28) | 59 (3.84) | 21 (2.28) |
| **Socioeconomic index (Mean, SD)** | -0.37 (1.03) | -0.04 (1.04) | 0.17 (1.02) |
| **Minimum dietary diversity (Mean, SD)** | 3.03 (1.42) | 3.35 (1.56) | 3.66 (1.62) |
| **Household food insecurity (Mean, SD)** | 1.67 (2.31) | 1.35 (2.07) | 1.32 (2.03) |

### LAZ Latent Class Trajectories by HAPIN Country Site and Sex

Individual latent class trajectories varied across countries between birth and 6 months of age, whereas trajectories were more consistent between 6 and 12 months of age. Children from all four HAPIN country sites had a mean LAZ below 0. Children in Peru had the highest mean LAZ at all time points followed by children in Rwanda. Children in India had the lowest mean LAZ at birth, across all four countries, but children in Guatemala had the lowest mean LAZ at all remaining timepoints. Notably, all countries exhibited decreases in growth trajectories between 6 and 12 months, although rates of decrease varied by country site (Figure 3).

**Figure 3.**
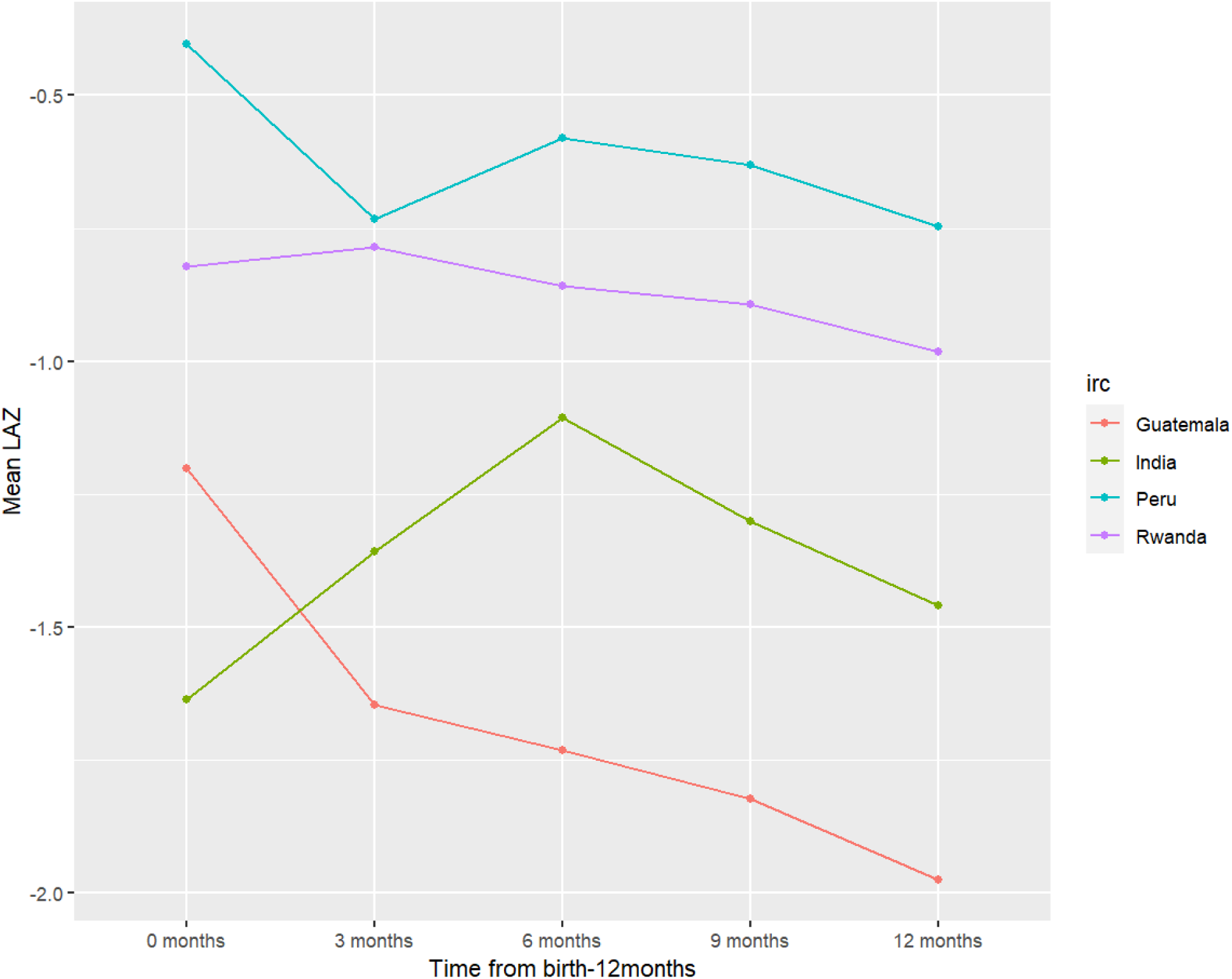
Mean length-for-age z-score (LAZ) by country site and across all timepoints

We used a multiple group latent class analysis to examine if patterns of linear growth vary as a function of HAPIN country (Model 2) or infant sex (Model 3) within the pooled sample and by infant sex within each HAPIN country site (Models 5-8). Model fit statistics for all subgroup analyses are presented in the Supplemental Materials (Table #S6-7). We did not identify any significant differences between latent class trajectories by country sites (p>0.9) (Model 2) or by sex (p>0.9) (Model 3). Additionally, there were no significant differences in latent growth trajectories by sex in three country samples, India (p=0.14), Peru (p=0.12) or Rwanda (p>0.9). However, we did identify significant differences in latent growth trajectories by sex specifically within the Guatemala sample (p<0.0001) (Model 5). Guatemalan boys had lower mean LAZ, respective to Guatemalan girls, in all trajectory groups and at all timepoints.

### LAZ Latent Class Trajectories by Timing of Intervention

Finally, we used a multiple group latent class analysis to examine if patterns of linear growth vary as a function of timing of intervention delivery during gestation. However, we identified no significant differences between those in the early and late gestation groups (p>0.9) (Model 4).

## DISCUSSION

In this study, we evaluated the impact of an LPG stove and free fuel intervention on linear growth trajectories during the first year of life in four country settings. We identified three distinct latent class trajectories, consistent with the literature on linear growth trajectories [9] [19] [34]. Despite high fidelity and adherence to the intervention and a large exposure contrast between intervention arms, we did not identify differences in latent class trajectories of LAZ growth between the intervention and control arms of the HAPIN trial [14, 16]. These results are consistent with previous analyses of fetal growth, birth weight, and stunting at 12 months in the HAPIN trial [8, 10, 35].

Our results on the differences in descriptive characteristics by class membership are consistent with the literature on known predictors of stunting. Maternal height and household wealth are known predictors of linear growth and are significantly associated with child stunting [36]. Preterm infants are underweight and stunted early in life, but they exhibit height and weight catch-up, primarily between 23-47 months of age [37, 38]. These collective results do not support the use of environmental-specific interventions to improve child growth outcomes. Further analyses are necessary to examine linear growth patterns beyond 12 months, and to assess if patterns beyond 12 months change as a result of postnatal household air pollution exposure from birth-12 months.

Our results differ from the prevailing observational literature that household air pollution exposure is associated with linear growth outcomes [7]. However, most of this literature relies on cross-sectional observational studies conducted in single countries, mostly in South Asia with some in sub-Saharan Africa. Most studies were primarily conducted in the postnatal period to examine the association of household air pollution exposure from solid fuel compared to cleaner fuel sources [7]. Observational household air pollution studies are also subject to uncontrolled confounding bias [39]. A few studies use longitudinal data and examine prenatal and postnatal exposure to household air pollution. The Ghana Randomized Air Pollution and Health Study (GRAPHS) examined the effect of both prenatal and postnatal household air pollution exposure on linear growth trajectories; similar to HAPIN, the cookstove intervention arm was not associated with linear growth trajectories [9]. However, GRAPHS reported that prenatal PM_2.5_ exposure was associated with impaired linear growth and increased stunting risk in the first year of life [9] based on a secondary exposure-response analysis with a limited number of participants in a single setting. And finally, the Randomized Exposure Study of Pollution Indoors and Respiratory Effect (RESPIRE) and follow-up Chronic Respiratory Effects of Early Childhood Exposure to Respirable PM (CRECER) cohort estimated that a 1ppm higher average carbon monoxide exposure was associated with a 0.21 lower height-for-age (HAZ), with the association being stronger among boys compared to girls [40]. Similarly to GRAPHS, the RESPIRE/CRECER intention-to-treat analysis indicated null results while the exposure-response indicated an effect on growth outcomes. Therefore, a next step will be to examine exposure-response relationships between PM_2.5_, BC and CO with patterns of LAZ growth in the HAPIN trial.

Interestingly, we only identified differences in latent growth trajectories by infant sex in Guatemala – for all latent class groups, the boys’ trajectory was consistently lower than the girls’ trajectory. These results are consistent with the literature on sex differences in linear growth – girls consistently have higher HAZ than boys [40–42]. However, it is unclear why these differences were only apparent in Guatemala. One explanation for the observed difference, specific to Guatemala, could be due to cultural perceptions of gender that result in different feeding behaviors. An ethnographic study conducted in an indigenous village in Guatemala highlighted that mothers reported introducing complementary foods early to infant boys because they perceived that breastmilk alone was not sufficient for infant boys [43]. Early introduction of complementary foods can lead to reduced consumption of breastmilk and its health-promoting factors which can increase exposure to illnesses that impair linear growth [43, 44].

Finally, we did not identify differences in individual latent class trajectories by timing of the installation of the LPG stove in HAPIN households. Infants born to women who received the intervention at less than 18 weeks’ gestation (33.8g, 95% CI −2.6 to 70.2) had a slightly greater birth weight compared to infants born to women who received the intervention after 18 weeks’ gestations (5.3g, 95% CI −31.0 to 41.7g) [10]. However, both GRAPHS and HAPIN imply that the timing of the deployment of the intervention may not have been sufficient to address prenatal risk factors that can adversely affect birth and growth outcomes, including linear growth. GRAPHS identified sensitive windows in girls at 10 weeks gestation and in boys from 16-20 weeks where prenatal carbon monoxide exposure was inversely associated with birth weight [45]. The heterogeneity in the exposure reduction between the intervention and control arms in GRAPHS also suggest that reduction in household air pollution exposure must occur *prior* to the sensitive window periods [46]. While mechanisms underlying the timing of intervention delivery to linear growth or stunting outcomes are currently unclear, it has been proposed that first trimester interventions can have the largest effect on fetal growth ratio and birth length, and subsequently influence linear growth status and stunting [47, 48]. Less than 2% (1.98%, n=28) of intervention mothers received the intervention in the first trimester (0-12 weeks) and thus, our results support earlier hypotheses in HAPIN that timing of intervention delivery beyond the first trimester may not affect linear growth trajectories.

Strengths of this study include the rigorous design, involving an RCT that was conducted in four geographically diverse sites with nearly exclusive adherence to the intervention and low attrition (10.8-13.5%) [8, 14, 16]. In addition, this analysis used high-quality repeated anthropometric measurements collected at ≥3 timepoints from over 2,800 children between birth and 12 months of age. And finally, latent class methods facilitated the identification of latent subgroups of growth by country site and timing of intervention, as well as to account for missing data using maximum likelihood under the missing at random assumption.

However, we acknowledge some limitations of this work. Firstly, there were interruptions to anthropometry data collection due to COVID-19 restrictions. As previously described, there were notable differences between intervention and control children with missing measurements during the period of COVID-19 restrictions [8]. For example, intervention children who were missing the length measurement at 12 months were born to mothers who were younger and taller and lived in households with less food insecurity than control children who were also missing the length measurement at 12 months [8].

Secondly, some participants have missing data; we did not use multiple imputation, similar to the primary HAPIN outcome papers [10]. However, we did use the maximum likelihood estimator in Mplus with the assumption that missing data were missing at random. And finally, although we were able to identify distinct classes of heterogenous latent growth in our data, latent class analysis assumes fixed variances within class and does not allow for within-class variation.

Our study offers a comprehensive examination of child linear growth trajectories between birth and 12 months of age, a critical period that is sensitive to prenatal exposures and associated with growth and development beyond the first year of life. This analysis uses the most recent longitudinal data, from multiple time points that were collected in a standardized manner and from multiple global regions, to allow examination of child linear growth using both pooled data and from each HAPIN country site. Our results corroborate the current school of thought from gold standard household air pollution - that an LPG cookstove intervention delivered after the first trimester is not independently sufficient to improve child linear growth outcomes between birth and 12 months of age.

## Data Availability

Data described in the manuscript, code book, and analytic code will be made available upon request.

## SUPPLEMENTAL TABLES

**Table #S1.** Missing data for length-for-age z-score score measurements by timepoint, country site and intervention arm assignment for 2802 children whose mothers’ completed participation in the HAPIN Trial.

|  | Number (%) of missing LAZ measurements |  |  |  |  |
| --- | --- | --- | --- | --- | --- |
|  | Birth | 3 Months | 6 Months | 9 Months | 12 Months |
| **Total sample size at each time point** | 2528 | 2093 | 2026 | 2036 | 2267 |
| **Percent (%) missing at each time point** | 10.84% | 13.09% | 13.52% | 13.46% | 12.09% |
| **Percent (%) missing by country site:** |  |  |  |  |  |
| **Guatemala** | 2.65% | 3.20% | 3.31% | 3.29% | 2.96% |
| **India** | 4.43% | 5.35% | 5.53% | 5.50% | 4.94% |
| **Peru** | 0.47% | 0.57% | 0.59% | 0.59% | 0.53% |
| **Rwanda** | 3.28% | 3.97% | 4.10% | 4.08% | 3.66% |
| **Percent (%) missing by intervention arm assignment:** |  |  |  |  |  |
| **Intervention** | 5.78% | 6.98% | 7.21% | 7.17% | 6.44% |
| **Control** | 5.06% | 6.12% | 6.32% | 6.29% | 5.65% |
| **Percent (%) missing by child sex:** |  |  |  |  |  |
| **Female** | 4.91% | 5.92% | 6.12% | 6.09% | 5.47% |
| **Male** | 5.93% | 7.17% | 7.40% | 7.37% | 6.62% |

**Table #S2.** Reasons for withdrawal or exiting the HAPIN Trial.

|  |
| --- |
| 135 mothers exited the HAPIN Trial before child's birth as a result of:<br>• 68 mothers experienced pregnancy loss<br>• 41 mothers voluntarily withdrew<br>• 21 mothers moved out from the catchment area<br>• 4 mothers were withdrawn by the study team<br>• 1 mother passed away |
| 110 children exited the HAPIN Trial before study completion due to:<br>• 58 child deaths<br>• 26 children moved from catchment area<br>• 20 children were voluntarily withdrawn<br>• 1 child was lost to follow-up |

**Table #S3.** Descriptive characteristics by HAPIN country site for 2802 children whose mothers’ completed participation in the HAPIN Trial.

|  | **Guatemala** | **India** | **Peru** | **Rwanda** |
| --- | --- | --- | --- | --- |
| **Maternal height at baseline (Mean, SD)** | 148.43 (5.31) | 151.17 (5.67) | 152.67 (4.48) | 156.58 (5.95) |
| **Maternal age at baseline (Mean, SD)** | 24.76 (4.43) | 24.00 (3.77) | 25.57 (4.54) | 27.36 (4.38) |
| **Gestational age at baseline (Mean, SD)** | 14.28 (3.04) | 16.07 (2.97) | 15.77 (3.35) | 15.50 (2.79) |
| **Nulliparity (Yes) (N, %)** | 209 (28.63%) | 407 (56.61%) | 229 (36.35%) | 205 (28.35%) |
| **Infant sex (Female) (N, %)** | 350 (47.95%) | 327 (45.48%) | 320 (50.79%) | 354 (48.96%) |
| **Preterm birth (Yes) (N, %)** | 41 (5.62%) | 42 (5.84%) | 25 (3.97%) | 28 (3.87%) |
| **Number of people living in household (Mean, SD)** | 5.18 (2.58) | 3.81 (1.55) | 4.55 (1.73) | 3.50 (1.44) |
| **Socioeconomic index (Mean, SD)** | -0.44 (0.73) | 0.97 (0.67) | 0.46 (0.60) | -0.96 (0.86) |
| **Minimum dietary diversity (Mean, SD)** | 3.05 (1.27) | 2.87 (1.13) | 5.03 (1.31) | 2.92 (1.51) |
| **Household food insecurity (Mean, SD)** | 1.17 (1.80) | 0.49 (1.20) | 1.38 (1.91) | 2.50 (2.65) |
| **Length-for-age z-score at six-months (Mean, SD)** | -1.73 (1.04) | -1.11 (1.18) | -0.580 (0.984) | -0.862 (1.13) |

**Table #S4.** Mean length-for-age z-score at each timepoint by intervention assignment and latent class trajectory for 2802 children whose mothers’ completed participation in the HAPIN Trial.

|  | **Intervention Low** | **Intervention Medium** | **Intervention High** | **Control Low** | **Control Medium** | **Control High** |
| --- | --- | --- | --- | --- | --- | --- |
| Birth | -2.27 | -1.15 | -0.19 | -2.42 | -1.30 | -0.42 |
| 3 Months | -2.79 | -1.31 | -0.12 | -2.95 | -1.42 | -0.28 |
| 6 Months | -2.69 | -1.36 | -0.01 | -2.91 | -1.41 | -0.12 |
| 9 Months | -2.78 | -1.51 | -0.08 | -3.02 | -1.54 | -0.21 |
| 12 Months | -2.92 | -1.65 | -0.26 | -3.10 | -1.66 | -0.41 |

**Table #S5.**
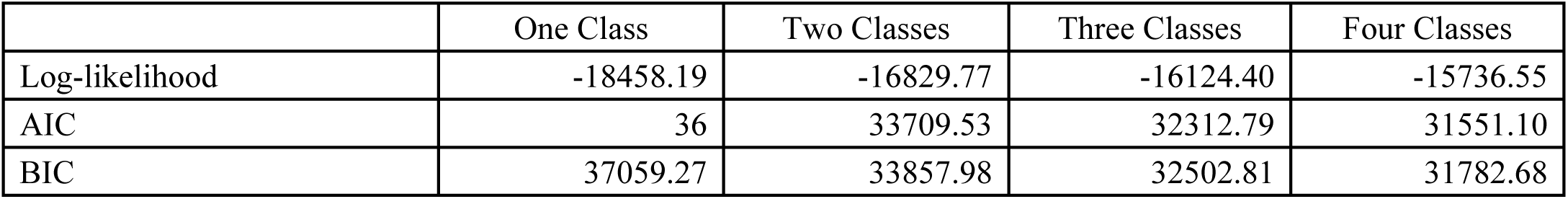
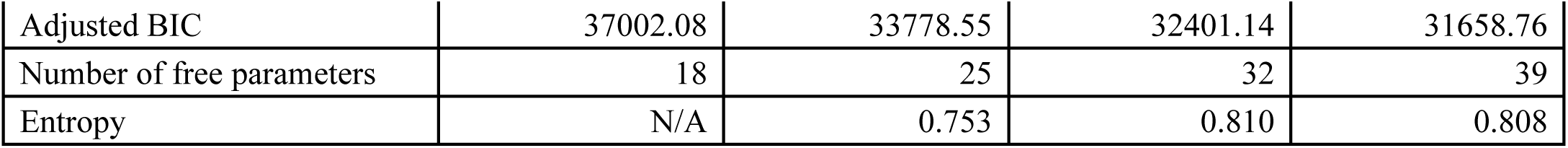
Model fit statistics for latent class trajectories of length-for-age z-score by intervention arm for 2802 children whose mothers’ completed participation in the HAPIN Trial.

|  | **One Class** | **Two Classes** | **Three Classes** | **Four Classes** |
| --- | --- | --- | --- | --- |
| Log-likelihood | -18458.19 | -16829.77 | -16124.40 | -15736.55 |
| AIC | 36 | 33709.53 | 32312.79 | 31551.10 |
| BIC | 37059.27 | 33857.98 | 32502.81 | 31782.68 |
| Adjusted BIC | 37002.08 | 33778.55 | 32401.14 | 31658.76 |
| Number of free parameters | 18 | 25 | 32 | 39 |
| Entropy | N/A | 0.753 | 0.810 | 0.808 |

**Table #S6.** Model fit statistics for latent class trajectories of length-for-age z-score by country site, sex, timing of intervention and by sex within each country site.

|  | HAPIN Country | Infant Sex | Timing of Intervention |
| --- | --- | --- | --- |
| Log-likelihood | -17791.34 | -16074.45 | -8091.96 |
| AIC | 35680.68 | 32206.89 | 16241.92 |
| BIC | 35971.65 | 32379.10 | 16394.25 |
| aBIC | 35815.96 | 32286.96 | 16302.13 |
| Number of free parameters | 49 | 29 | 29 |
| Entropy | 0.809 | 0.798 | 0.79 |

|  | Guatemala | India | Peru | Rwanda |
| --- | --- | --- | --- | --- |
| Log-likelihood | -4145.982 | -4130.948 | -3170.245 | -4186.355 |
| AIC | 8343.963 | 8313.896 | 6392.489 | 8424.71 |
| BIC | 8463.382 | 8432.92 | 6508.078 | 8543.878 |
| aBIC | 8380.824 | 8350.363 | 6425.531 | 8461.321 |
| Number of free parameters | 26 | 26 | 26 | 26 |
| Entropy | 0.827 | 0.707 | 0.787 | 0.859 |

\*Akaike Information Criterion (AIC) and Bayesian Information Criterion (BIC) are methods for scoring and selecting a model. AIC puts more emphasis on model performance and selects more complex models whereas BIC penalizes complex models. The Adjusted Bayesian Information Criterion (aBIC) is the sample size-adjusted BIC.

## Acknowledgements

The HAPIN trial is funded by the US National Institutes of Health (NIH; cooperative agreement 1UM1HL134590) in collaboration with the Bill & Melinda Gates Foundation (OPP113127). A multidisciplinary, independent data and safety monitoring board (DSMB) appointed by the US National Heart, Lung, and Blood Institute (NHLBI) monitors the quality of the data and protects the safety of patients enrolled in the HAPIN trial. NHLBI DSMB: Catherine J Karr (Chair), Nancy R Cook, Stephen Hecht, Joseph Millum, Nalini Sathiakumar (deceased), Paul K Whelton, Gail Weinmann, and Thomas Croxton (Executive Secretary). Programme coordination: Gail Rodgers, Bill & Melinda Gates Foundation; Claudia L Thompson, National Institute of Environmental Health Sciences; Mark J Parascandola, US National Cancer Institute; Marion Koso-Thomas, Eunice Kennedy Shriver National Institute of Child Health and Human Development; Joshua P Rosenthal, Fogarty International Center; Concepcion R Nierras, NIH Office of Strategic Coordination, The Common Fund; Katherine Kavounis, Dong-Yun Kim, Antonello Punturieri, and Barry S Schmetter (deceased), NHLBI. The findings and conclusions in this report are those of the authors and do not necessarily represent the official position of the NIH or US Department of Health and Human Services. This research represents the NIH’s contribution to the Global Alliance for Chronic Diseases (GACD) coordinated call for research on prevention and management of chronic lung diseases for 2016. We also thank Patrick Breysse, Donna Spiegelman, and Joel Kaufman (members of the advisory committee) for their valuable insight and guidance throughout the implementation of the trial. We also wish to acknowledge all the research staff and study participants for their dedication to and participation in this important trial.

Graduate student support was provided by the Laney Graduate School at Emory University and the National Institute of Environmental Health (Award Number 5T32ES12870). We are grateful to Usha Ramakrishnan for supervision support and Ghislaine Rosa, Stella M. Hartinger and Marilu Chiang for support with the trial implementation.

---

**Supplemental Figure 1.**
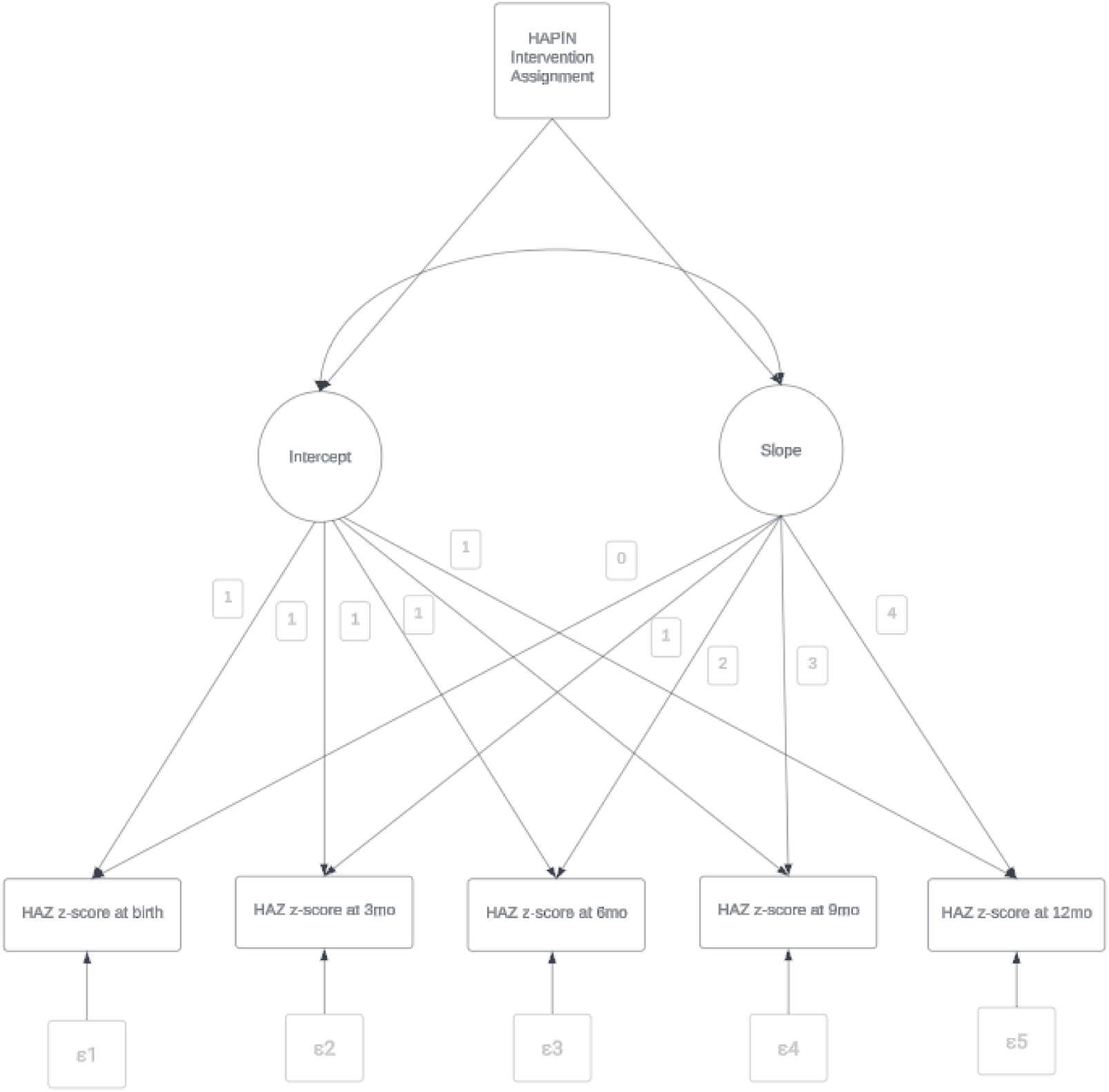
Path diagram of a longitudinal latent growth curve model. Observed variables are represented by squares and latent variables are represented by circles, with residuals represented by the small circles with an error term. The arrows between the latent variables, the intercept and slope, and the observed variables are fixed in advance. The factor loadings are fixed to 1 for the intercept latent variable and the factor loadings for the slope latent variable are fixed according to the change in time.

**Supplemental Figure 2.**
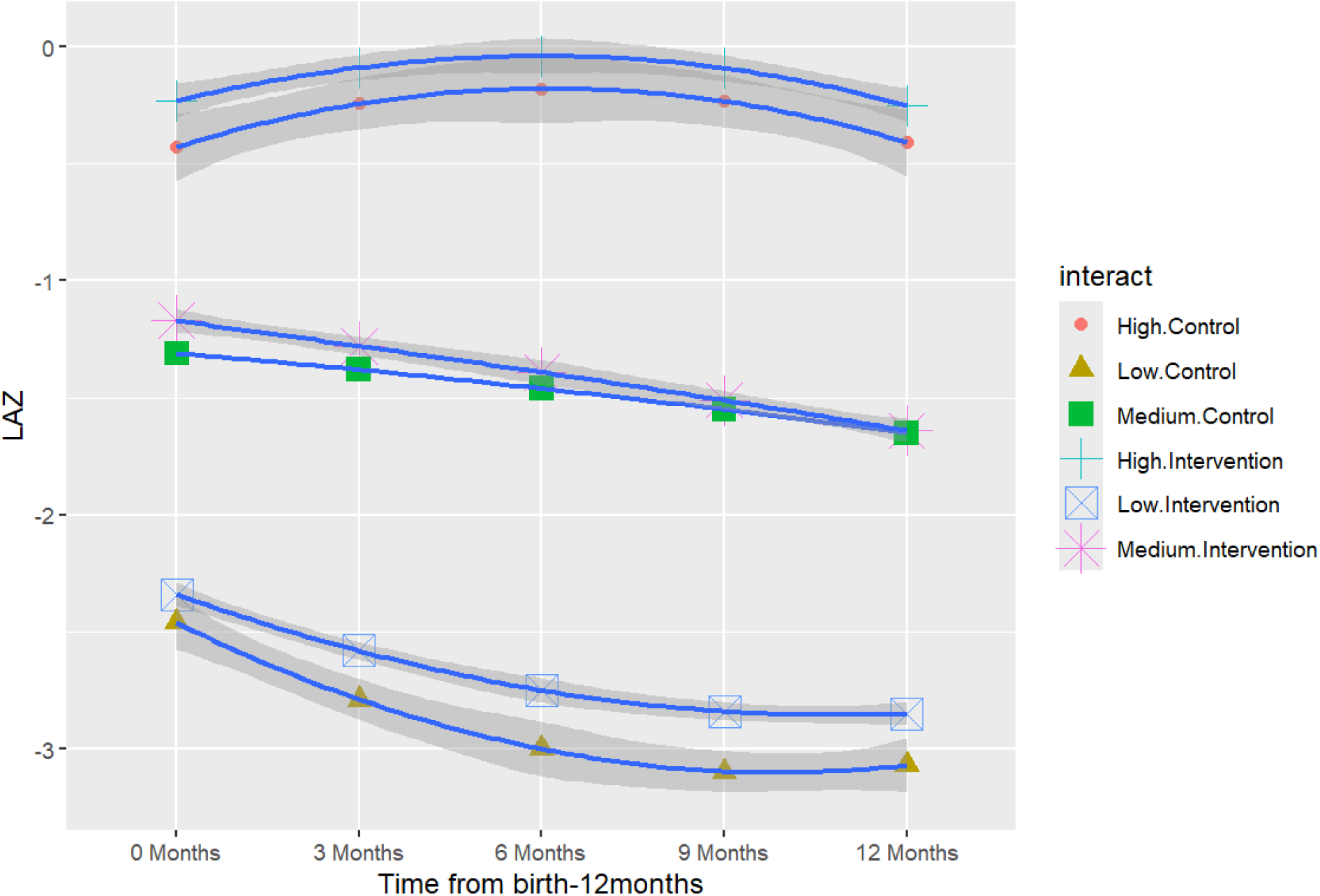
Latent classes by intervention arm for intention-to-treat analysis

**Supplemental Figure 3.**
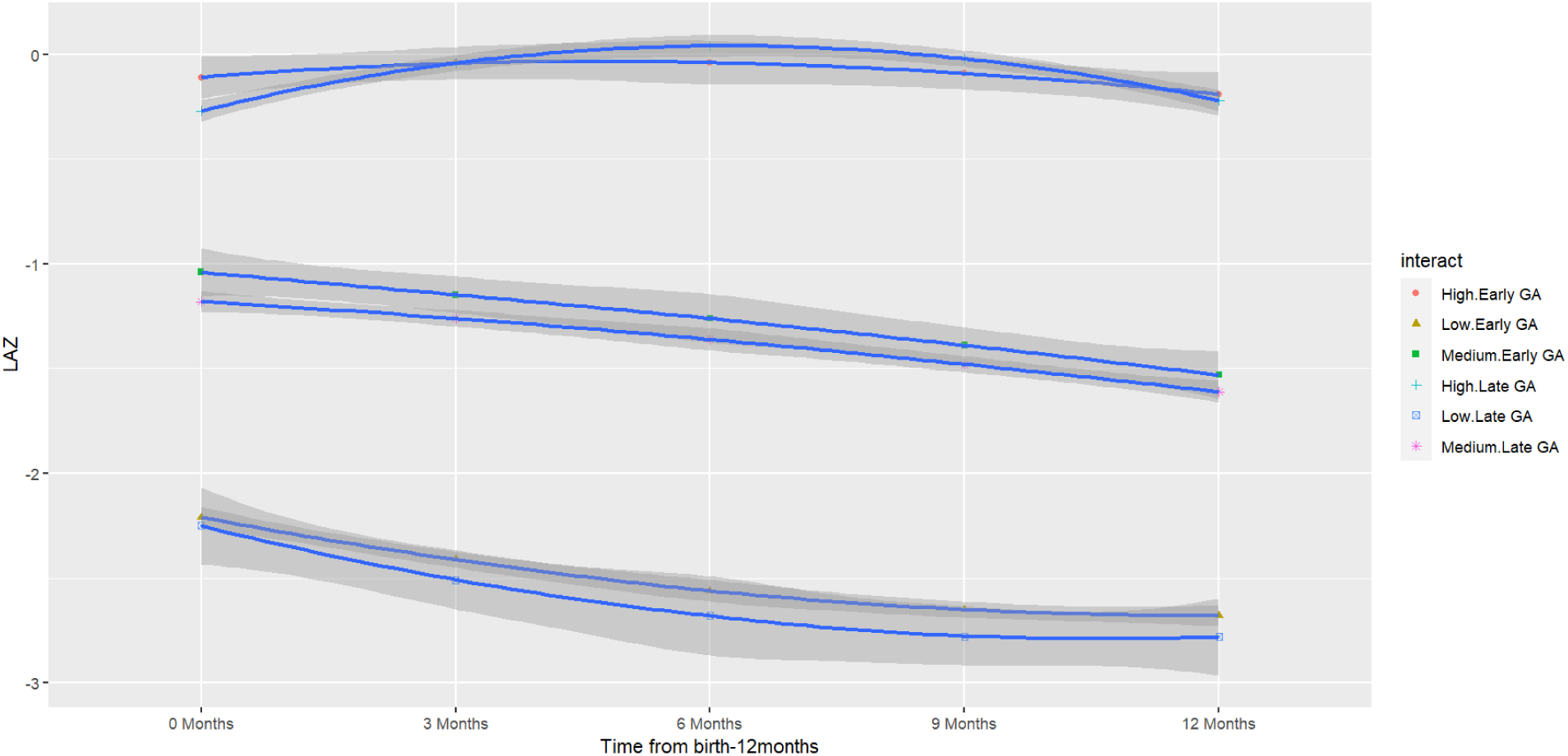
Latent classes for timing of intervention delivery analysis

**Supplemental Figure 4.**
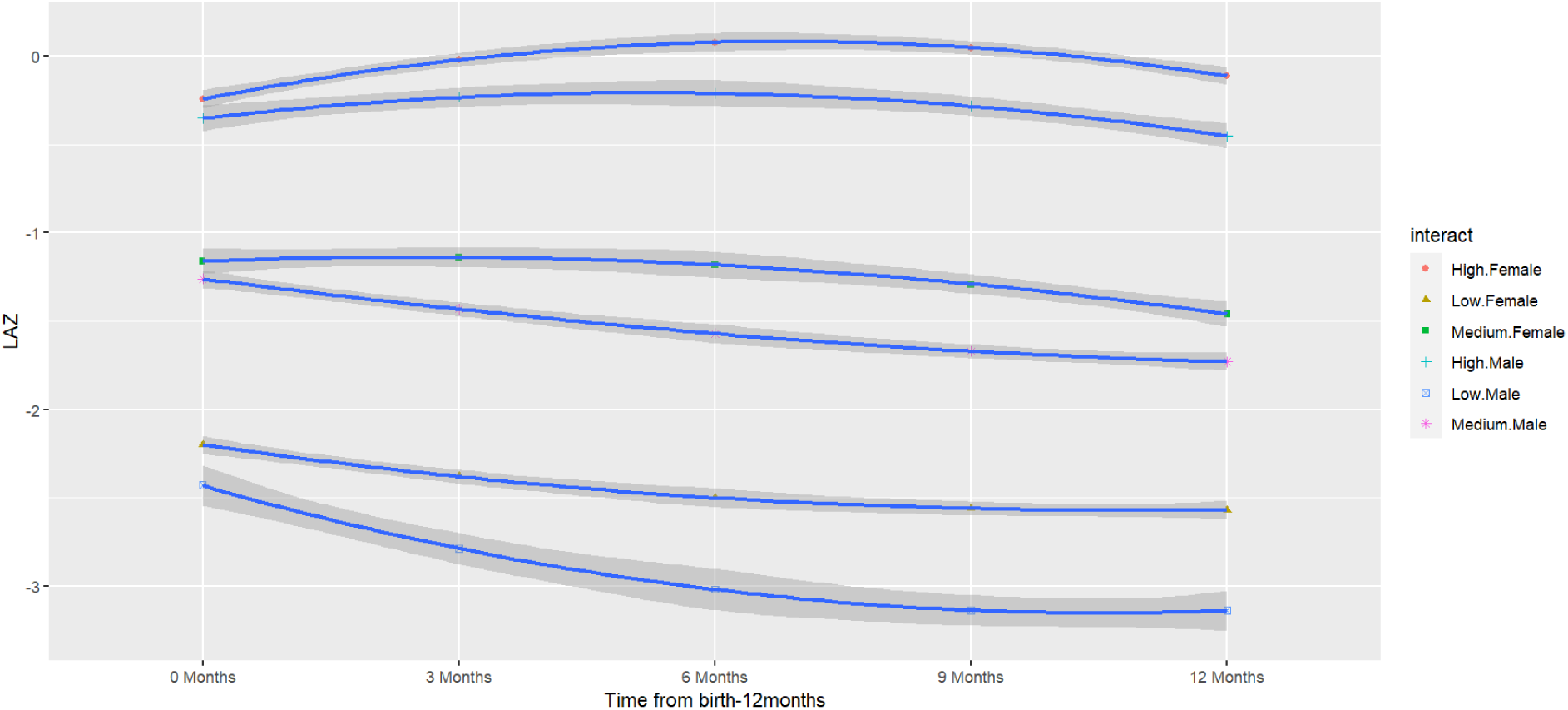
Latent classes for sex analysis within Guatemala sample

